# Portable Breath-Based Volatile Organic Compound Monitoring for the Detection of COVID-19: Challenges of Emerging Variants

**DOI:** 10.1101/2022.09.06.22279649

**Authors:** Ruchi Sharma, Wenzhe Zang, Ali Tabartehfarahani, Andres Lam, Xiaheng Huang, Anjali D. Sivakumar, Chandrakalavathi Thota, Shuo Yang, Robert P. Dickson, Michael W. Sjoding, Erin Bisco, Carmen Colmenero Mahmood, Kristen Machado Diaz, Nicholas Sautter, Sardar Ansari, Kevin R. Ward, Xudong Fan

**Affiliations:** Department of Biomedical Engineering, University of Michigan, Ann Arbor, MI, U.S.A.; Max Harry Weil Institute for Critical Care Research and Innovation, University of Michigan, Ann Arbor, MI, U.S.A.; Department of Electrical Engineering and Computer Science, University of Michigan, Ann Arbor, MI, U.S.A.; Department of Emergency Medicine, University of Michigan, Ann Arbor, MI, U.S.A.; Department of Internal Medicine, Division of Pulmonary Critical Care Medicine, University of Michigan, Ann Arbor, MI, U.S.A.

**Author notes:** Equal contribution (Ruchi Sharma, Wenzhe Zang).

## Abstract

**Importance:** Breath analysis has been explored as a non-invasive means to detect COVID-19. However, the impact of the emerging variants such as Omicron on the exhaled breath profile and hence the accuracy of breath analysis is unknown.

**Objective:** To evaluate the diagnostic accuracies of breath analysis on detecting COVID-19 patients in periods where Delta and Omicron were most prevalent.

**Design, Setting, and Participants:** A convenience cohort of patients testing positive and negative for COVID-19 using reverse transcriptase polymerase chain reaction (RT-PCR) were studied and included 167 COVID and non-COVID patients recruited between April 2021 and May 2022, which covers the period when Delta (and other variants prior to Delta) was the dominant variant (April – December 2021) and when Omicron was the dominant variant (January – May 2022). The breath from those patients were collected and analyzed for volatile organic compounds (VOCs) with a newly developed portable gas chromatography-based breath analyzer. Diagnostic patterns and algorithms were developed.

**Results:** A total of 205 breath samples were analyzed from 167 COVID and non-COVID patients. The RT-PCR was conducted within 18 hours of the breath analysis to confirm the COVID status of the patients. Among 94 COVID positive samples, 41 samples were collected from the patients in 2021 who were assumed to be infected by the Delta variant (or other variants occurring in 2021) and 53 samples from the patients in 2022 who were assumed to be infected by the Omicron variant (BA.1 and BA.2). Breath analysis using a set of 4 VOC biomarkers was able to distinguish between COVID (Delta and other variants in 2021) and non-COVID with an overall accuracy of 94.7%. However, the accuracy dropped significantly to 82.1% when the same set of biomarkers were applied to the Omicron variant with and 21 out of 53 COVID positive being misidentified. A new set of 4 VOC biomarkers were found to distinguish the Omicron variant and non-COVID, which yielded an overall accuracy of 90.9%. Breath analysis was also found to be able to distinguish between COVID (for all the variants occurring between April 2021 and May 2022) and non-COVID with an overall accuracy of 90.2%, and between the Omicron variant and the earlier variants (Delta and other variants occurring in 2021) with an overall accuracy of 91.5%.

**Conclusions and Relevance:** Breath analysis of VOCs using point of care gas chromatography may be a promising diagnostic modality for detection of COVID and similar diseases that result in VOC production. However, similar to other diagnostic modalities such as rapid antigen testing, challenges are posed by the dynamic emergence of viral variants. The results of this study warrant additional investment and evaluation on how to overcome these challenges and to exploit breath analysis to improve the diagnosis and care of patients.

**Key Points:** *Question:* Can volatile organic compounds (VOCs) in exhaled breath provide diagnostic information on COVID-19? Will variants such as Omicron B.1.1.529 and others affect the accuracy in breath analysis?

*Findings:* A set of 4 VOC biomarkers were found to distinguish between Delta (and the variants occurring in 2021) from non-COVID. The Omicron variant (occurring in 2022) significantly affects VOC profiles requiring the search for a new set of VOC biomarkers to distinguish between Omicron and non-COVID.

*Meanin:* These findings demonstrate the ability of breath analysis to distinguish between COVID and non-COVID, but also reveal the significant difference in the exhaled breath profile between COVID-19 patients during the period when Delta was most prevalent and when Omicron was most prevalent.

## 1. Introduction

The ongoing COVID-19 pandemic continues to present diagnostic challenges due to emerging variants. The two major platforms for COVID-19 diagnosis continue to be reverse transcriptase polymerase chain reaction (RT-PCR) testing from deep nasal swabbing and rapid antigen testing (RAT) from samples obtained from more proximal nasal swabbing. While RT-PCR is the gold standard, RAT was developed to address the need for faster turnaround as well as widespread scaling of testing to the public for those both asymptomatic and symptomatic. However, the transition of COVID-19 from the Delta variant to the Omicron variant has been demonstrated to significantly reduce the accuracy of RATs, thus presenting significant challenges in their ability to maintain accuracy which are critical to decision making^1-4^.

Since the beginning of the pandemic, alternative methods of detection of COVID-19 infection have been and continue to be explored, including the use of exhaled breath using a number of technologies such as GC-IMS (gas chromatography in tandem with ion mobility spectrometry), Fourier-transform infrared spectroscopy, GC-MS (mass spectrometry), and others^5-20^. The basis for these approaches is that exhaled breath contains hundreds of volatile organic compounds (VOCs), many of which are produced in response to inflammation and infection^19,21-36^. Several of these exhaled breath technologies have been demonstrated to have accuracies comparable to RT-PCR and RATs, and one has recently been approved for use under the FDA’s Emergency Use Authorization (EUA) pathway^10,17,18,37,38^.

However, nearly all results reported in these studies were prior to 2022 in which the dominant COVID-19 strain was the Delta variant. Here we report the use of a portable gas chromatography (GC) device developed as a point-of-care diagnostic for COVID-19 and its performance during the COVID-19 Delta surge and its transition to Omicron, including future challenges in using breath analysis in the current pandemic as well as future respiratory based pandemics.

## 2. Methods

### 2.1 Clinical study protocol and participants

This project was developed under the National Institutes of Health’s Screening for COVID-19 by Electronic-Nose Technology (SCENT) program in response to the declared public health emergency issued by the Department of Health and Human Services (DHHS) for the 2019 Novel Coronavirus (COVID-19) as part of the Rapid Acceleration of Diagnostics-Radical (RADx-rad) initiative^39^. The study was approved by the University of Michigan Medical School’s Institutional Review Board (HUM00103401 and HUM00210064). Informed consent from patients or their legally authorized representative was required.

Patients were enrolled between April 26, 2021 and May 31, 2022 through the adult Emergency Department at the University of Michigan Health System (Michigan Medicine). Michigan Medicine is a quaternary care center, and its Emergency Department provides care to over 70,000 adult patients per year. Convenience sampling was used to enroll patients undergoing RT-PCR testing who were symptomatic for COVID-19 or who were admitted for any presentation but required RT-PCR COVID-19 testing to ensure proper inpatient cohorting if they were found to be positive. Samples for RT-PCR tests were collected using nasopharyngeal swabbing for the patients who could breathe spontaneously and using either nasopharyngeal swabbing or tracheal aspirate for those who were on mechanical ventilators. Michigan Medicine utilizes a number of RT-PCR platforms including DiaSorin Molecular Simplexa™ Abbott ID NOW, Abbott Alinity, and Biofire FilmArray Respiratory Panel 2.1 to detect SARS-CoV-2 and other virus infections. No attempt was made to control for which RT-PCR test was utilized.

### 2.2. Exhaled breath collection and analysis

In the present study, we recruited patients who either were on a ventilator or could breathe spontaneously. Breath collection/analysis was conducted within 18 hours of the corresponding RT-PCR test described above. For mechanically ventilated patients, breath was collected into a 5 L Tedlar bag through a T-connector attached to the ventilator’s expiratory port, as shown in Figure 1(A). A portable pump was used to assist breath collection. Patients who could breathe spontaneously were asked to orally exhale into a 5 L Tedlar bag via a mouthpiece and a medical-grade HEPA filter (see Figure 1(B)) with the HEPA filter serving to prevent viral contamination of the Tedlar bag. Figure S1 provides greater visual details on the two breath sampling modes. Approximately 1 L of breath was collected for portable GC analysis.

**Figure 1.**
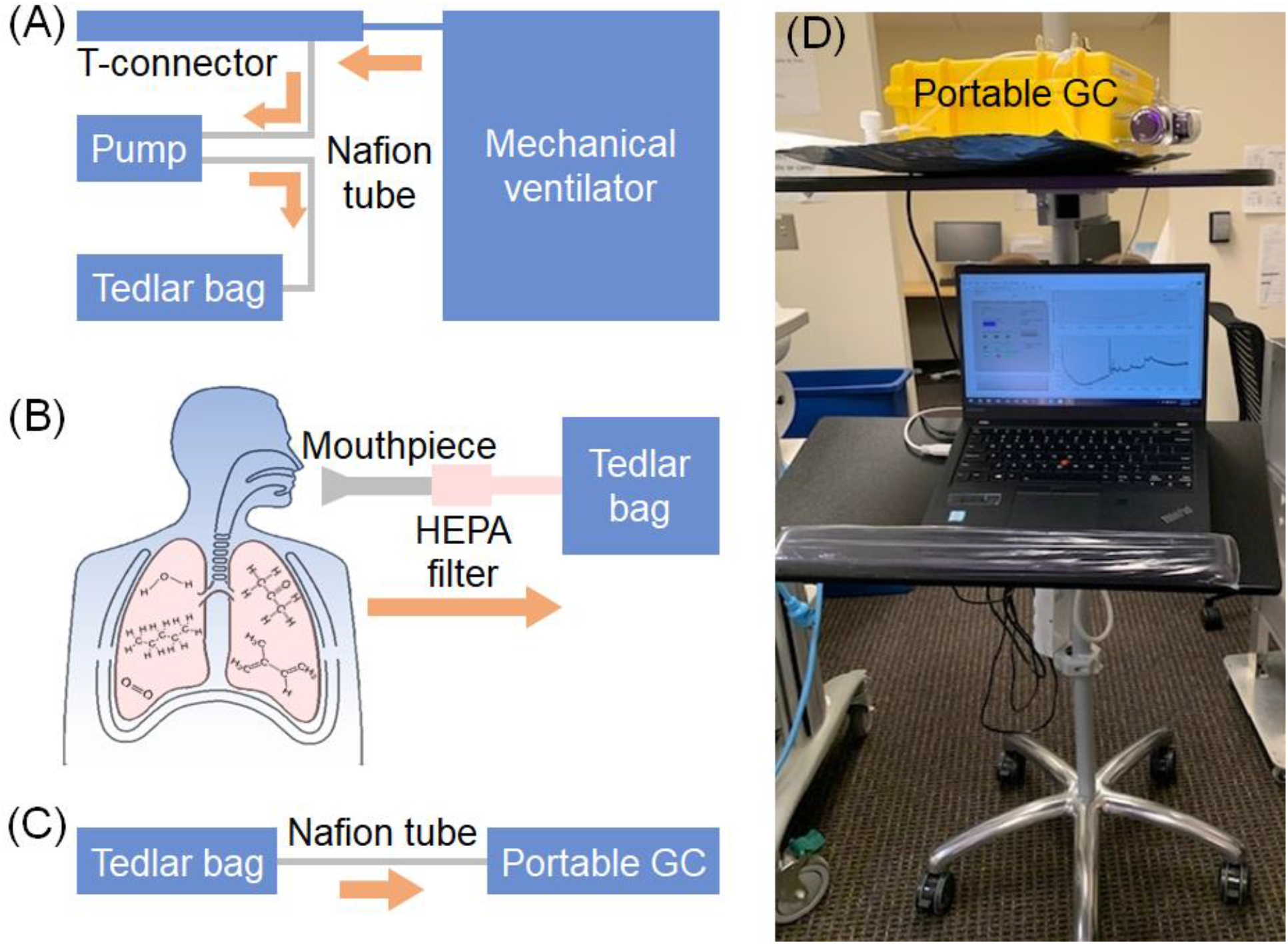
Two breath sampling modes. **(A)** For a patient on a ventilator, the breath sample was collected into a 5 L Tedlar bag via a T-connector connected to the expiratory port on the ventilator. Nafion tubes, which remove the moisture in the breath, were used to connect the T-connector, the pump, and the Tedlar bag. Details of the connection to the ventilator are shown in Figure S1. **(B)** For other patients who could breathe spontaneously, the breath sample was collected via a mouthpiece and a high efficiency particulate absorbing (HEPA) filter - the same filter as used in a ventilator to efficiently remove viruses, bacterial, and aerosols. **(C)** The breath sample in a Tedlar bag was subsequently sampled into and analyzed by the automated portable GC device that could be remotely controlled. **(D)** Photo showing a portable GC (housed in a yellow box) secured on a mobile cart, as well as a tethered laptop for on-board control.

Breath analysis took place in the patient’s room immediately after breath collection for all patients. The Tedlar bag was connected to the sampling port of the portable GC device via a Nafion filter (Figures 1(C) and (D)). A total of ∼350 mL of breath was sampled from the Tedlar bag into the portable GC at a rate of ∼70 mL/min for 5 minutes. The operation of the GC device can be found in Section S1 in the Supplementary Information. The details of the GC device itself have been described previously^21^. The entire process takes 25 minutes, including 10 minutes of sampling and internal sample transfer, 10 minutes of chromatographic separation, and 5 minutes of final internal cleaning. The GC operation was controlled remotely via a tethered laptop (Figure 1(D)) through the in-house developed LabVIEW™ codes.

Once the breath analysis was complete, the consumables including T-connector, Tedlar bag, mouthpiece, and HEPA filter were disposed of. All other parts such as the portable pump, Nafion tube, GC device, laptop, and mobile cart were pulled out of the patient’s room after disinfection using Oxy-bleach wipes.

### 2.3. Data analysis and biomarker discovery

After GC chromatogram signal pre-processing (such as baseline correction and de-noising), approximately 90 peaks could be detected in each breath chromatogram (Figure 2). Collectively, there were a total of 131 different peaks in the 205 chromatograms. Each peak represents one VOC or one set of co-eluted VOCs. The normalized peak area is used to quantify the relative amount of VOC(s) in each breath sample.

**Figure 2.**
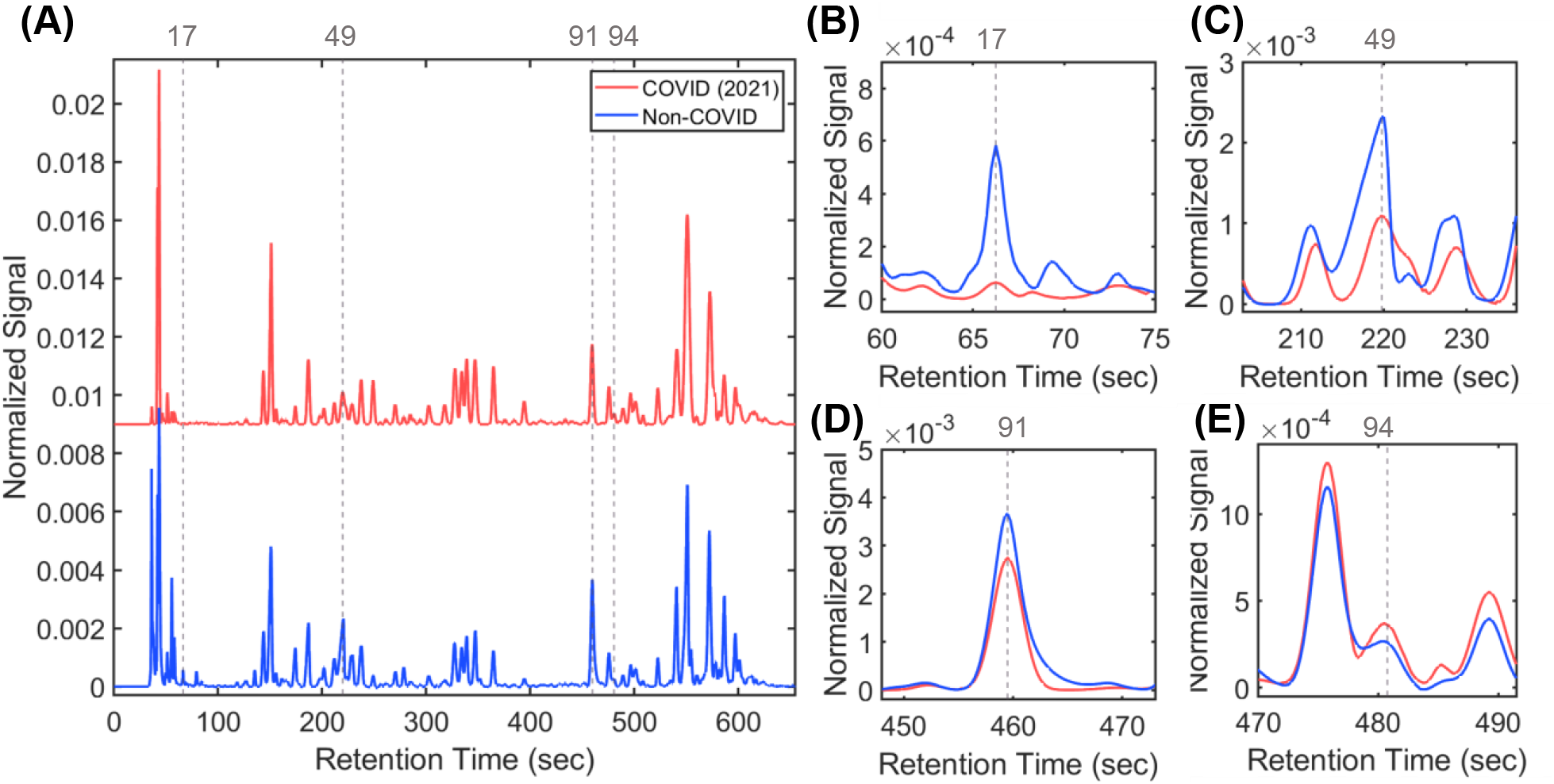
**(A)** Representative chromatograms of breath samples from a COVID patient collected in early July of 2021 and a non-COVID patient. The vertical grey dashed lines and numbers show the positions and listing numbers of the VOC biomarkers that distinguish between COVID (2021) and non-COVID. The details of these VOC biomarkers are given in Table 1. The violin charts of these VOCs for both COVID (2021) and non-COVID are given in Figure S4. **(B-E)** Zoom-in portions of **(A)**. Note that each chromatogram is normalized to the total area under the curve (from 0 s to 650 s). The COVID (2021) chromatogram is vertically shifted for clarity. COVID (2021) patients refer to those who were recruited prior to December 14, 2021 before the Omicron outbreak in the United States, and were therefore assumed to be infected by Delta or earlier SARS-CoV-2 variants (after April 2021). Non-COVID patients refer to those who were recruited throughout the study (from April 26, 2021 to May 31, 2022).

Finally, machine learning, linear discriminant analysis (LDA), and principal component analysis (PCA) were used for reducing the dimensionality of the datasets, biomarker selection for classification, and statistical analyses. LDA and PCA were initially chosen because of our past experience with this approach, which has been tested and validated in our earlier breath analysis studies on other diseases^21-23,40,41^. Details of the biomarker discovery algorithm were previously reported^21^. The primary analytic outcome was binary (to distinguish between COVID-19 positive versus negative when compared to RT-PCR).

**Table 1.** VOC biomarkers used in the study. (1) Biomarkers (Peak ID: 17, 49, 91, and 94) to distinguish between COVID (2021) and non-COVID. (2) Biomarkers (Peak ID: 17, 67, 87, and 97) to distinguish between COVID (2022) and non-COVID. (3) Biomarkers (Peak ID: 27, 67, 69, 87, and 94) to distinguish COVID (2021) and COVID (2022). (4) Biomarkers (Peak ID: 12, 67, 84, and 105) to distinguish between COVID positive (all COVID variants occurring between April 2021 and May 2022) and non-COVID. COVID (2021) refers to the COVID positive patients who were recruited prior to December 14, 2021, and were therefore assumed to be infected by Delta or earlier variants. COVID (2022) refers to the COVID positive patients who were recruited January 11 and May 31, 2022, and were therefore assumed to be infected by Omicron. Non-COVID refers to the COVID negative patients who were recruited throughout the study from April 26, 2021 to May 31, 2022. All recovered patients are also included in “non-COVID”. Note that the VOCs labeled with a “*” (such as Peak 12 and 67, *etc*.) appear in multiple biomarker sets.

| Peak ID | Compound Name |
| --- | --- |
| 12 | Methylcyclopentane |
| 17* | Benzene |
| 27 | Unidentified |
| 49 | Octane |
| 67* | 2,2,4-Trimethylheptane |
| 69 | 2,2-Dimethyloctane |
| 84 | Decene and Dimethyloctadiene (different isomers) |
| 87* | Decane |
| 91 | Trimethyloctane (different isomers) |
| 94* | Methyldecane (different isomers) |
| 97 | Undecane |
| 105 | Dimethyldecane and Tetramethyloctane (different isomers) |
| Biomarker set |  |
| COVID (2021) vs. non-COVID | 17*, 49, 91, 94* |
| COVID (2022) vs. non-COVID | 17*, 67*, 87*, 97 |
| COVID (2021) vs. COVID (2022) | 27, 67*, 69, 87*, 94* |
| COVID (all variants) vs. non-COVID | 12, 67*, 84, 105 |

### 2.4. VOC identification

In order to identify the chemical identities of the VOC biomarkers, we analyzed a subset of collected breath samples using both our portable GC and an Agilent mass spectrometer (MS). The outlet of the GC was connected to the MS via a 1 m long guard column as the transfer line (Figure S3(A)) so that the same breath samples could be detected simultaneously by the non-destructive photoionization detector (PID) on our GC and the MS. GC-MS is considered a gold standard to identify VOCs. See Section S2 in the Supplementary Information for details.

## 3. Results

The demographic details and the total number of subjects enrolled in this study are summarized in Figures 3(A)-(E). A total of 77 COVID patients and 91 non-COVID patients, as verified by the RT-PCR testing were recruited to obtain 205 breath samples. 26 subjects (11 COVID patients and 15 non-COVID patients) recruited in 2021 were collected/analyzed over multiple days (>24 hours interval) during their stay in the hospital for longitudinal monitoring. As per our IRB, we were allowed to collect breath samples up to ten days from the consent date. Figure 3(F) shows the transition from Delta to Omicron in Michigan^42^ between late December, 2021 and early January, 2022.

**Figure 3.**
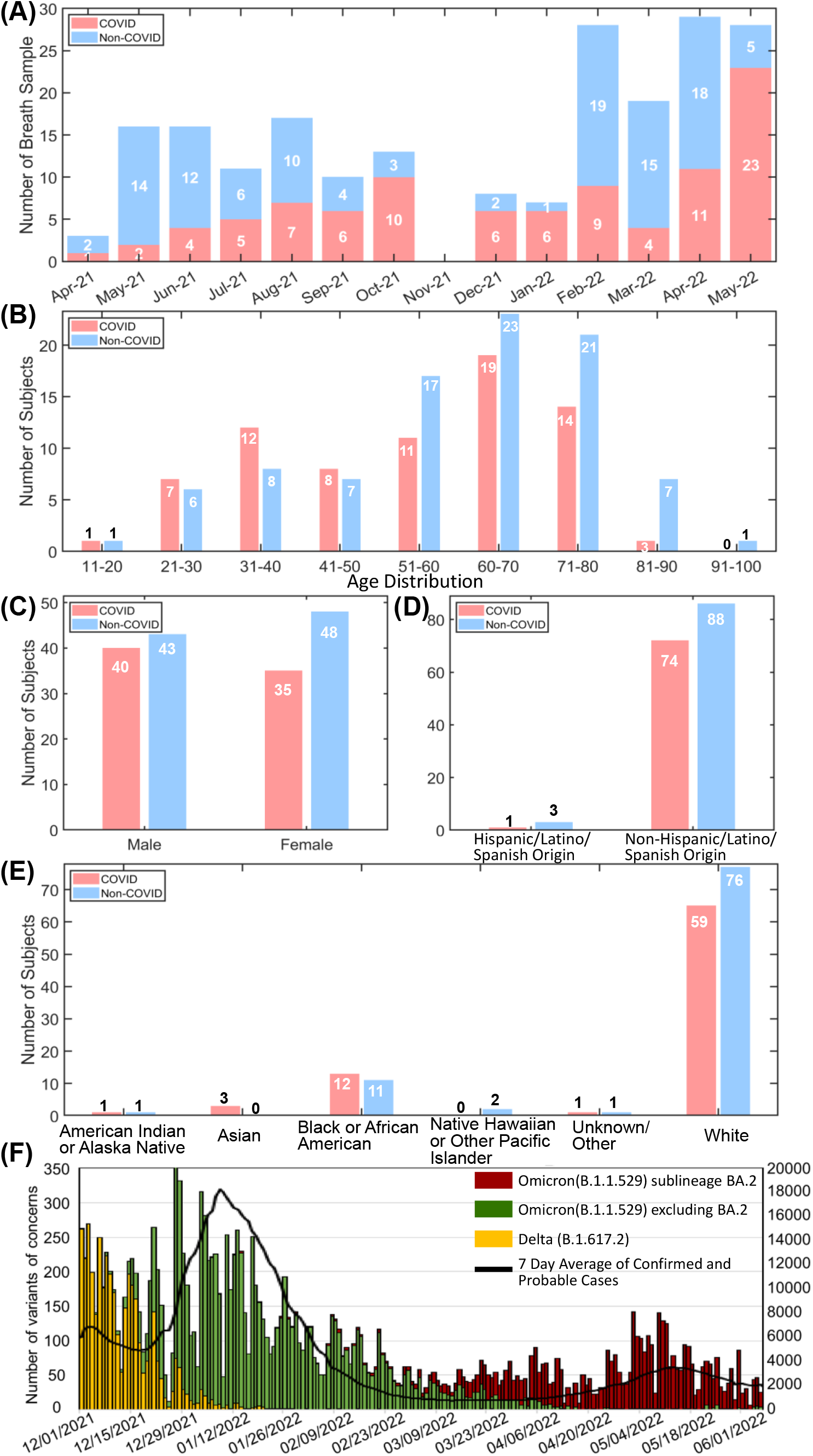
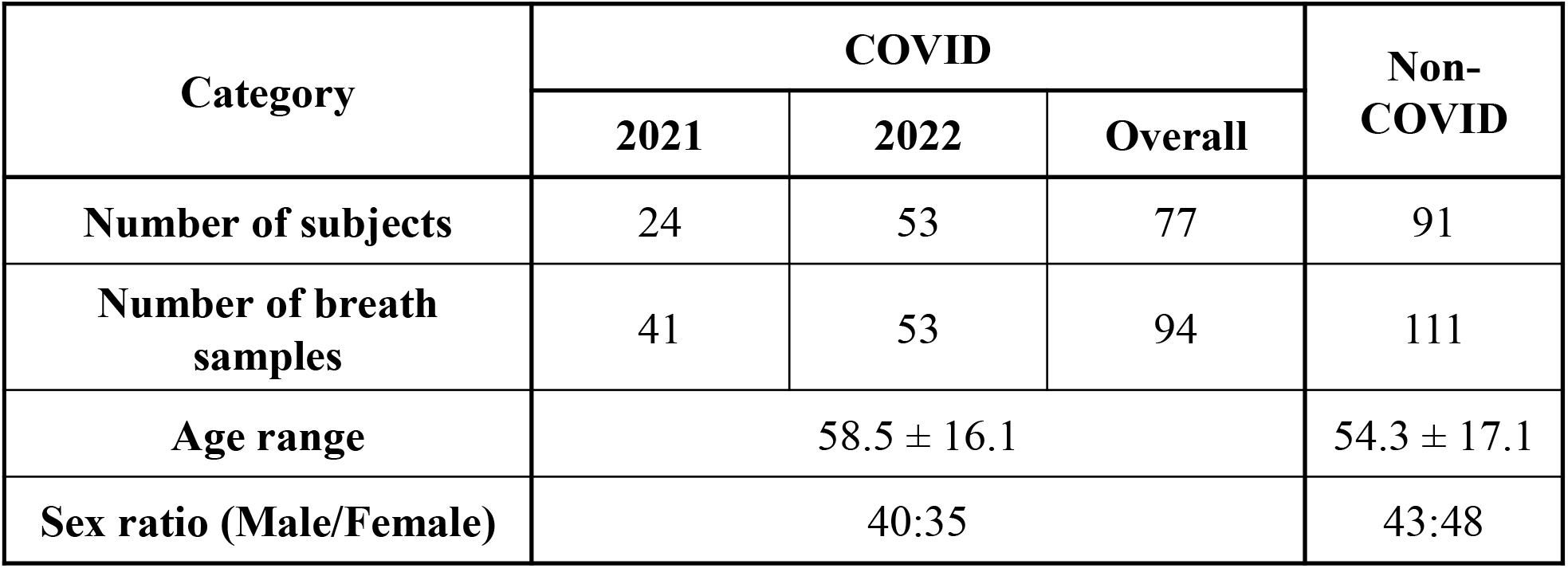
**(A-E)** Demographic information of the human subjects in this study. **(F)** shows the transition from Delta to Omicron in Michigan (adapted from Slide 7 in Ref. 42). **(Note 1)** The total number of subjects in this study is 167. One patient in 2021 was COVID positive and was later recovered (*i.e*., COVID negative). Therefore, this patient is counted in both COVID and non-COVID categories. **(Note 2)** In 2021, 26 subjects (11 COVID patients and 15 non-COVID patients) recruited were monitored for up to 10 days. Therefore, the number of breath samples is larger than the number of patients in 2021. **(Note 3)** In 2021, 2 COVID positive patients were asymptomatic. 3 breath samples from them were collected and analyzed. In 2022, 8 COVID positive patients were asymptomatic. 8 breath samples from them were collected and analyzed. **(Note 4)** The patients in 2021 were recruited between April 26 and December 14, 2021. The patients in 2022 were recruited between January 11 and May 31, 2022. **(Note 5)** No pregnant female patient was recruited in the study.

### 3.1. Distinguishing between COVID (2021) and non-COVID

Figure 2(A) shows a representative chromatogram of a breath sample from a COVID (2021) patient collected in early July of 2021 and a non-COVID patient. COVID (2021) patients refer to those recruited prior to December 14, 2021 and were therefore assumed to be infected by Delta or earlier variants^42,43^. Non-COVID patients refer to those who were recruited throughout the entire study (from April 26, 2021, to May 31, 2022). For VOC biomarker discovery, 48 out of 152 breath chromatograms were treated as the training set, among which 24 chromatograms were from 24 COVID positive patients, and 24 chromatograms were from 24 non-COVID patients. The remaining 104 chromatograms (17 COVID positive and 87 non-COVID) were used as the testing set.

Four VOCs (Peak ID # 17, 49, 91, and 94) were identified (see Figure 2 and Table 1) that were able to distinguish between COVID (2021) and non-COVID. A sensitivity of 92.7%, a specificity of 95.5%, a positive predictive value (PPV) of 88.4%, a native predictive value (NPV) of 97.2%, and an overall accuracy of 94.7% are achieved (Table 2), which provide performance for training, testing, combined training and testing. The results are equivalent to or higher than previously reported breath analysis results involving patients in 2020 and 2021^8-10,12-15,17,18,38^. The corresponding PCA plot is provided in Figure S5. Note that in Figure S5, there were 12 breath samples from 4 recovered patients who are correctly identified as non-COVID. Recovered patients refer to those who had been previously COVID positive (according to the RT-PCR tests) but were tested COVID negative at the time when the breath samples were collected and analyzed.

**Table 2.** Statistics summary corresponding to Figure S5 for COVID (2021) vs. non-COVID.

|  | Training |  |  | Testing |  |  | Training+Testing |  |  |
| --- | --- | --- | --- | --- | --- | --- | --- | --- | --- |
|  | COVID (2021) | Non-COVID | Total | COVID (2021) | Non-COVID | Total | COVID (2021) | Non-COVID | Total |
| **Positive** | 23 | 1 | 24 | 15 | 4 | 19 | 38 | 5 | 43 |
| **Negative** | 1 | 23 | 24 | 2 | 83 | 85 | 3 | 106 | 109 |
| **Column total** | 24 | 24 | 48 | 17 | 87 | 104 | 41 | 111 | 152 |
| **Specificity** | 95.8% |  |  | 95.4% |  |  | 95.5% |  |  |
| **Sensitivity** | 95.8% |  |  | 88.2% |  |  | 92.7% |  |  |
| **Positive predictive value** | 95.8% |  |  | 78.9% |  |  | 88.4% |  |  |
| **Negative predictive value** | 95.8% |  |  | 97.6% |  |  | 97.2% |  |  |
| **Total accuracy** | 95.8% |  |  | 94.2% |  |  | 94.7% |  |  |

### 3.2. Distinguishing between COVID (2022) and non-COVID

COVID (2022) patients refer to those recruited after January 11, 2022 and to the end of the study (May 31, 2022) and were therefore assumed to be infected by the Omicron variant^42-44^. There was a gap of nearly one month between our last recruitment in 2021 and the first recruitment in 2022. Using the four previously identified VOC biomarkers in Section 3.1 (*i.e*., Peak ID # 17, 49, 91, and 94), breath analyses on COVID (2022) resulted in a significantly lower sensitivity (60.4%) and accuracy (82.1%) but without a change in specificity (95.4%), since there is no change in the non-COVID cases in the testing set given in Table 2). This suggested a different pathologic response of humans to Omicron, as compared to Delta and earlier variants occurring in 2021 (denoted as COVID (2021)). Consequently, the same set of biomarkers of Delta and earlier variants appeared not to be suitable for Omicron (*i.e*., COVID (2022)). As such, a new VOC biomarker search was undertaken in the same manner as was performed for COVID (2021).

For the new VOC biomarker search, 50 out of 164 breath chromatograms were treated as the training set, among which 25 chromatograms were from 25 COVID positive patients and 25 chromatograms were from 25 non-COVID patients. The remaining 114 chromatograms (28 COVID positive and 86 non-COVID) were used as the testing set. A new set of VOC biomarkers (Peak ID # 17, 67, 87, and 97) were identified (see Figure S7 and Table 1) that were able to distinguish between COVID (2022) and non-COVID. A sensitivity of 88.7%, a specificity of 91.7%, a PPV of 83.9%, an NPV of 94.4%, and an overall accuracy of 90.9% are achieved (Table 3). The corresponding PCA plot is given in Figure S8.

**Table 3.** Statistics summary corresponding to Figure S8 for COVID (2022) vs. non-COVID.

|  | Training |  |  | Testing |  |  | Training+Testing |  |  |
| --- | --- | --- | --- | --- | --- | --- | --- | --- | --- |
|  | COVID (2022) | Non-COVID | Total | COVID (2022) | Non-COVID | Total | COVID (2022) | Non-COVID | Total |
| **Positive** | 23 | 1 | 24 | 24 | 8 | 32 | 47 | 9 | 56 |
| **Negative** | 2 | 24 | 26 | 4 | 78 | 82 | 6 | 102 | 108 |
| **Column total** | 25 | 25 | 50 | 28 | 86 | 114 | 53 | 111 | 164 |
| **Specificity** | 96.0% |  |  | 90.7% |  |  | 91.9% |  |  |
| **Sensitivity** | 92.0% |  |  | 85.7% |  |  | 88.7% |  |  |
| **Positive predictive value** | 95.8% |  |  | 75.0% |  |  | 83.9% |  |  |
| **Negative predictive value** | 92.3% |  |  | 95.1% |  |  | 94.4% |  |  |
| **Total accuracy** | 94.0% |  |  | 89.5% |  |  | 90.9% |  |  |

### 3.3. Distinguishing between COVID (2021) and COVID (2022)

Since the pathology and human response are different between Delta (and earlier variants) and Omicron, it may be possible to distinguish them using breath analysis. In the new round of biomarker search, we used 48 out of 94 breath chromatograms as the training set, among which 24 were from 24 COVID (2021) patients and 24 were from 24 COVID (2022) patients. The remaining 46 chromatograms (17 COVID (2021) and 29 COVID (2022) were used for testing.

A new set of VOC biomarkers (Peak ID # 27, 67, 69, 87, and 94) were identified (see Figure S7 and Table 1) that were able to distinguish between presumed Omicron and all previous variants. A sensitivity of 92.7%, a specificity of 90.6%, a PPV of 88.4%, an NPV of 94.1%, and an overall accuracy of 91.5% are achieved (Table 4). The corresponding PCA plot is given in Figure S9.

**Table 4.** Statistics summary corresponding to Figure S9 for COVID (2021) vs. COVID (2022).

|  | Training |  |  | Testing |  |  | Training+Testing |  |  |
| --- | --- | --- | --- | --- | --- | --- | --- | --- | --- |
|  | COVID (2021) | COVID (2022) | Total | COVID (2021) | COVID (2022) | Total | COVID (2021) | COVID (2022) | Total |
| **Positive** | 22 | 2 | 24 | 16 | 3 | 19 | 38 | 5 | 43 |
| **Negative** | 2 | 22 | 24 | 1 | 26 | 27 | 3 | 48 | 51 |
| **Column total** | 24 | 24 | 48 | 17 | 29 | 46 | 41 | 53 | 94 |
| **Specificity** | 91.7% |  |  | 89.7% |  |  | 90.6% |  |  |
| **Sensitivity** | 91.7% |  |  | 94.1% |  |  | 92.7% |  |  |
| **Positive predictive value** | 91.7% |  |  | 84.2% |  |  | 88.4% |  |  |
| **Negative predictive value** | 91.7% |  |  | 96.3% |  |  | 94.1% |  |  |
| **Total accuracy** | 91.7% |  |  | 91.3% |  |  | 91.5% |  |  |

### 3.4. Distinguishing between COVID (all variants) and non-COVID

An additional question and concern is whether breath analysis can distinguish between COVID positive and non-COVID regardless of variants. Practically, a set of VOC biomarkers that are suitable for all variants (*i.e*., the variants occurring between April 2021 and May 2022, including Delta, Omicron BA.1, and Omicron BA.2) would be very useful for rapid COVID diagnostics without considering its variant. Towards this end, we conducted another round of biomarker search by using 96 out of 205 breath chromatograms as the training set, among which 48 were from 48 COVID positive patients (24 COVID (2021) patients and 24 COVID (2022) patients)) and 48 were from 48 non-COVID patients. The remaining 109 chromatograms (46 COVID positive and 63 non-COVID) are used as the testing set, including those collected in May of 2022.

A new set of VOC biomarkers (Peak ID # 12, 67, 84, and 105) were identified (see Figure S7 and Table 1) that were able to distinguish between COVID (all variants occurring between April 2021 and May 2022) and non-COVID. A sensitivity of 89.4%, a specificity of 91.0%, a PPV of 89.4%, an NPV of 91.0%, and an overall accuracy of 90.2% are achieved (Table 5). The corresponding PCA plot is given in Figure S10.

**Table 5.**
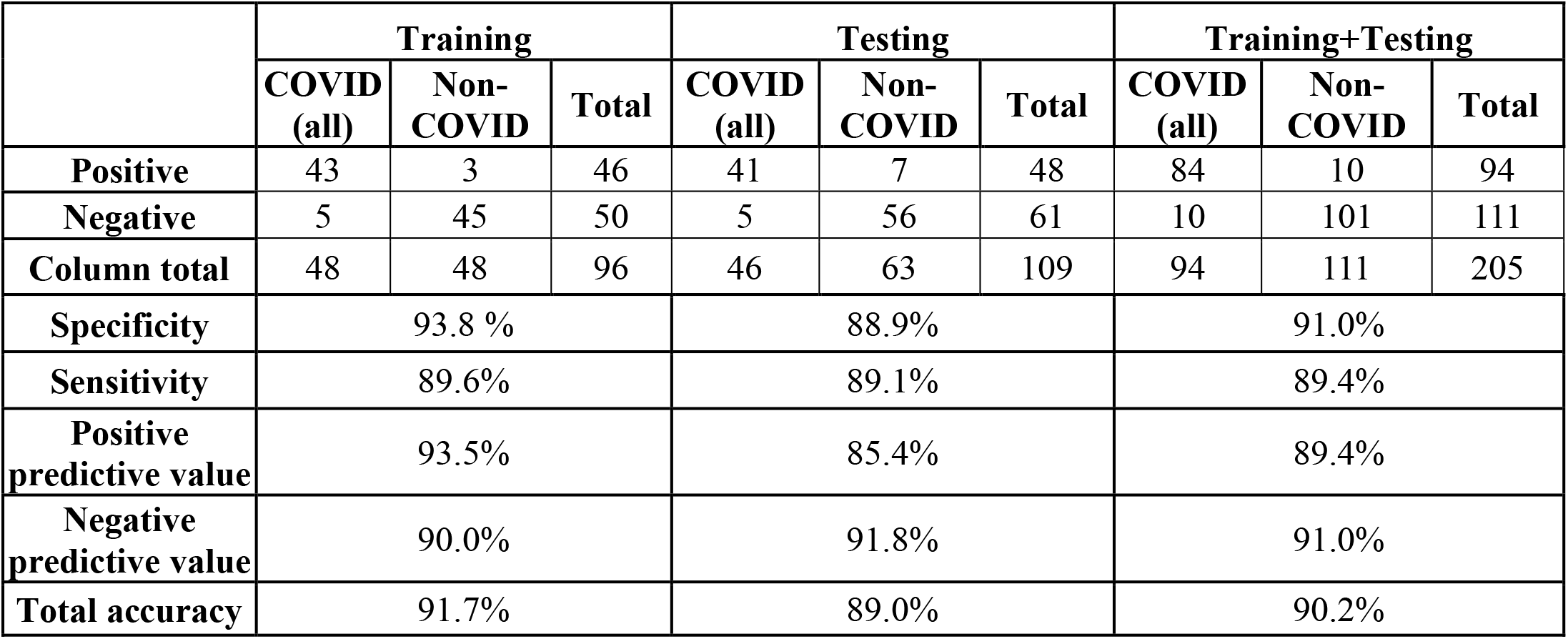
Statistics summary corresponding to Figure S10 for COVID (all variants occurring between April 2021 and May 2022) vs. non-COVID.

## 4. Discussion

In this study, we demonstrated the ability of a portable GC technology to detect exhaled breath VOC signatures at point-of-care that were capable of diagnosing COVID-19 with accuracies that could potentially make it a candidate diagnostic platform. More importantly, we also demonstrated the challenge that emerging variants pose to such technologies. On November 30, 2021, the COVID-19 Omicron variant B.1.1.529 was determined to be a variant of concern by the U.S. and since has (with additional subvariants) become the dominant COVID-19 strain in the U.S. During our study, we noted a significant decrease in sensitivity (from 88.2% to 60.4%) beginning in January, 2022. It was during this time that the Omicron variant began to become the dominant COVID-19 strain. While Michigan Medicine does not do subtyping of the COVID-19 virus, we relied on CDC’s reports as well as surveillance and subtyping information provided by the state of Michigan’s Department of Health^42,43^. Figure 3(F) provides the data from the State of Michigan during the 2021-2022 transition period.

Since the VOCs detected in our study are likely reflective of inflammation, this may not be surprising, because the Delta variant was associated with more severe inflammation^45-47^. As the COVID-19 virus has mutated over time, these mutations have impacted viral characteristics such as transmissibility and symptom severity. Unlike the ability of RT-PCR that relies on conserved RNA, these mutations have significantly impacted the performance of RATs and other molecular tests^48^. Other confounding variables that are difficult to account for that could affect VOC signatures related to inflammation include changes in the basic biology of the COVID-19, rates of vaccination and boosters, and the advent of outpatient treatments such as Paxlovid™, *etc*.

As indicated in the Results section, a new VOC biomarker search was performed once we noted the drop in performance and the national and regional reporting of the rapid emergence of the new variant, which significantly improved the accuracy of breath analysis. These VOC combinations appear to be able to distinguish patterns different from those observed during the peak of the Delta variant. When the entire pattern library and models were combined, the overall performance on all subjects attained accuracies that could potentially make breath analysis a viable and rapid testing alternative. This mainly becomes of great importance during the transition of one variant to another to ensure that declines in diagnostic accuracies are short lived as variants can emerge at different rates of speed. For implementation of such a strategy great vigilance will need to occur and require careful monitoring and communication with health authorities and their subtyping efforts in order to allow for new VOC signature identification that can then be incorporated to ensure accurate diagnosis regardless of the variant during the time of variant transition.

While we used RT-PCR as a gold standard, there continues to be debated regarding its accuracy and concerns over false negative, which have been reported to be between 1% and 30%^1,49-51^. False negatives can occur for many reasons ranging from testing that occurs too early or late, poor sample collection, changes in viral shedding, and others. As such, our performance could potentially be better or with further study could be used as a confirmatory strategy or as possible adjunct to study and determine risk for transmissibility or the development of complications linked to inflammation severity. Indeed, we observed 6 subjects who tested both positive and negative with RT-PCR within several hours of each other due to duplicative testing orders being placed. These subjects were removed from our analysis.

As demonstrated in our previous studies of acute respiratory distress syndrome (ARDS) we were able to note the evolution of severe COVID and its trajectory ^22,23^. In the current study, we were able to monitor some COVID positive patients (all recruited in 2021) for up to 10 days.

Section S3 in the Supplementary Information provides information on 5 patients to highlight the potential of breath analysis in monitoring COVID patients’ trajectories and predicting their clinical outcomes. The cases include following some patients who recovered as well as those who deteriorated. As such, a potential advantage of breath for patients with severe disease resulting in respiratory failure and requiring mechanical ventilation, could be the ability to continuously monitor them.

During our study, we were also able to capture a subset of asymptomatic COVID patients and non-COVID patients who were also found to be infected by other viruses such as rhinovirus, human metapneumo virus, human coronavirus OC43 (HCoV-OC43), and enterovirus. The results for those patients are discussed in Section S4 in the Supplementary Information. The presence of these viruses did not appear to significantly confound the results of GC testing, but further studies are needed.

As compared to other technologies such as GC-IMS, GC-MS, and Fourier-transform infrared spectrometry, the portable GC device presented here is much lighter in weight and smaller in size. The high-performance photo-ionization detector (PID) detector used allows for the detection of VOCs in tens of ppt (parts-per-trillion) in 350 mL of breath sample^52^. Recently, the first FDA EUA using breath analysis on COVID detection was issued to InspectIR^37^, which is a GC-MS, and larger and heavier (∼20 kg) GC-MS than our device (∼3 kg).

There are several limitations and challenges to our study and the use of GC as a diagnostic modality in the setting of COVID and other future viral pandemics. We did not perform COVID subtyping but instead relied on our state’s and the CDC’s reports of the incidence of emerging variants^42,43^. Currently we are examining our data collected before the Omicron BA.4 and BA.5 variant. We expect to experience decrease accuracy requiring a new search for diagnostic breath VOC signatures. This is a potential major disadvantage of breath analysis technology but will require more advanced analytic work to identify a universal signature that is perhaps more conserved and less prone to change. Similar to RATs, there will need to be an agreed upon regulatory approach as to how to track and report the performance of breath diagnostics as variants emerge.

We also do not clearly understand the etiology of the VOCs produced during COVID infection but assume based on their identification are related to the process of inflammation that may be occurring in both the upper and lower respiratory tract^22,23,31,53^. The fact that we were able to diagnose COVID in both symptomatic and asymptomatic subjects is encouraging. More studies on these particular VOCs including their origins may assist in the development of a better understanding of the disease as well as the potential to develop new diagnostics.

Lastly, we used a limited data science approach to identifying diagnostic VOC patterns including the use of LDA and PCA. Additional machine learning tools such the use of Random Forrest and Neural Networks may prove to increase the diagnostic accuracy of the approaches. More advanced features selection techniques such as LASSO (least absolute shrinkage and selection operator) regression and Minimum Redundancy Maximum Relevance (mRMR) may result in more accurate and stable selection of VOCs. Finally, the current data analysis pipeline involves several manual steps such as peak alignment. Efforts are underway to create a fully automated pipeline using a combination of convolutional neural networks and constrained integer optimization techniques.

## 5. Conclusion

Breath analysis of VOCs using portable gas chromatography at point of care may be promising diagnostic modality for detection of COVID and like diseases that result in VOC production. However, similar to other diagnostic modalities such as rapid antigen testing, challenges are posed by the dynamic emergence of viral variants. The results of this study warrant additional investment and evaluation of how to overcome these challenges and to exploit breath analysis to improve the diagnosis and care of patients.

## Data Availability

All data produced in the present study are available upon reasonable request to the authors

## Acknowledgments

The authors acknowledge the support from the National Institutes of Health via U18TR003812 and the Office of the Director of National Intelligence (ODNI), Intelligence Advanced Research Projects Activity (IARPA), via IARPA FA8650-19-C-9101. The views and conclusions contained herein are those of the authors and should not be interpreted as necessarily representing the official policies or endorsements, either expressed or implied, of the ODNI, IARPA, or the U.S. Government. The U.S. Government is authorized to reproduce and distribute reprints for Governmental purposes notwithstanding any copyright annotation thereon. The authors acknowledge microfabrication aid from the Lurie Nanofabrication Facility at the University of Michigan. The authors also acknowledge the support of the Max Harry Weil Institute for Critical Care Research and Innovation including its Proposal Development Unit, Clinical Research Unit and Data Science Unit.

