## Supplementary Information for "Portable Breath-Based Volatile Organic Compound Monitoring for the Detection of COVID-19: Challenges of Emerging Variants"

*: Equal contribution

### S1. Breath sampling, GC operation, and characterization

S1.1. Breath sampling

The breath sample was collected into a 5 L Tedlar bag as shown in Figure S1 at a flow rate of 70 mL/min for 5 minutes.

S1.2. GC operation

The details about the portable GC itself can be found in our previous study[1]. The operation of the portable GC is described briefly:

1. Connection of the Tedlar bag to the GC inlet;
2. Withdrawal of the breath sample into the GC device by GC internal pump;
3. GC separation of the breath sample
4. GC self-cleaning

The total time for Steps 1-4 is ~25 minutes.

S1.3. Inter-GC characterization

Five portable GC devices were constructed for this study. To evaluate the repeatability, the same breath sample collected from a non-COVID subject was respectively analyzed by those GC devices. In addition, a chromatogram of normal alkanes mixtures, $C_6$-$C_{11}$, is obtained from one of the five GC devices operated under identical conditions (such as temperature ramping and flow rate, *etc*.) to mark the breath VOC peak positions against those of $C_6$-$C_{11}$ (Figure S2). It is seen that all breath VOC peaks are aligned well (within +/- 0.5 seconds) after correlation optimized warping algorithms, which allows us to pinpoint the peaks among the chromatograms obtained from different GC devices by using their respective retention times. All the chromatograms (and the subsequent processed data) in the study were obtained from those five GC devices. In our data analysis, we treat all data equally regardless of the GC device used to obtain them.

For each individual breath sample, the number of chromatographic peaks range is approximately 90. The total number of chromatographic peaks among all patients who we recruited in the study is 131. We label them from Peak 1 to Peak 131 from the earliest peak to the latest peak.

### S2. GC-MS identification of breath biomarkers

To chemically identify the breath biomarkers, the outlet of our portable GC was connected to an Agilent mass spectrometry (MS), as illustrated in Figure S3(A), which allows us to compare the chromatograms obtained concomitantly by the photoionization detector (PID) in our GC and MS for the same breath sample (see Figure S3(B)).

GC-MS is gold-standard for VOC identification in complex matrices, but this method can fail when the obtained data are contaminated with additional molecule fragments due to coelution. These additional fragments can lead to VOC misidentification by automated MS software, due to the reduced the spectrum match factor below the identification threshold and the presence of contaminated fragments. In order to address this problem, we developed the following pipeline for more accurate identification of breath biomarkers.

1. Use the NIST MS library for chemical identification. The NIST MS library often provides many possible “hits”.
2. The MS spectrum and mass fragments are manually checked to confirm the presence of the main mass fragments in the NIST suggested compound by MS library.
3. Check whether the suggested compound in Step 1 is present in human breath from previous studies. Particularly, the website maintained by EPA (https://comptox.epa.gov/dashboard/chemical_lists/VOLATILOME) is adopted, which provides a list of 1117 compounds (as of November 2021) in human breath.
4. Further narrow down the compound candidates by comparing their vapor pressure (or boiling point) with their neighboring normal alkanes (Figure S2).

The final identification of the breath biomarkers is listed in Table 1 in the main text.

### S3. Trajectory monitoring of COVID (2021) patients

One of the prominent advantages of breath analysis is its ability of continuous and non-invasive monitoring of patients, as demonstrated in our previous studies[1-3]. In the current study, some COVID positive patients (all recruited in 2021) were monitored for up to 10 days since their recruitment into the study. Below we present 5 cases to highlight the potential of breath analysis in monitoring COVID patients' trajectories and predicting their clinical outcomes. Such ability was also demonstrated in our previous studies in acute respiratory distress syndrome in both human and swine[2,3].

***Recovery cases.*** Figures S11(A) and (B) show the trajectories of four COVID patients (Patients *a*, *b*, *c*, and *d*). All the patients were initially COVID positive, and later recovered and were discharged from the hospital.

Patient *a* was sampled for 6 days (Day 2, 3, 4, 9, and 10). Until Day 4, the patient was COVID positive. On the 9$^{th}$ day the patient was listed as non-COVID based on the RT-PCR within 18 hours of breath analysis. Later this patient got extubated and discharged from the hospital, approximately 2 months after our last day of breath analysis.

Patient *b* on Day 2 (data point marked as *b*2) represents the most severe case that we saw among the entire patient pool, whose data point is the farthest in the distance from the boundary (grey line in the PCA plot). As per our GC measurement, this patient showed recovery (moving towards the gray boundary line) as we monitored the patient breath on Day 2, 4, and 8. We could not continue our measurement further after the patient consent expired, but later this patient recovered and was discharged from the hospital 15 days after our last breath analysis. We observed a similar trajectory with Patients *c* and *d*. Their breath measurements showed the recovery trend (moving towards the gray boundary line); later those two patients recovered and were discharged from the hospital after last breath measurement.

***Deterioration case***. Patient *e* was COVID positive, and his/her situation deteriorated over time. This patient died 21 days after our last breath measurement (on Day 10). Figure S11(C) shows the patient got worsened with continuous breath measurement (the data point moving farther away from the gray boundary line on the PCA plot).

### S4. Asymptomatic patients and cross-reactivity examination

During our study, we also closely monitored the asymptomatic COVID patients and non-COVID patients infected by other viruses such as rhinovirus, human metapneumo virus, HCoV-OC43, and enterovirus. For better clarity, their breath analysis data are re-plotted in Figures S12-S14 using the same PCA plot as in Figures S5, S8, and S9, respectively.

Below we provide a list of symptoms used to screen symptomatic and asymptomatic COVID patients. (1) We first examined the COVID RT-PCR test; (2) then examined the upper respiratory infection (URI) symptoms (cough, sore throat, and shortness of breath); (3) looked for other signs known to be related to COVID such as abdominal pain, nausea, vomiting, diarrhea, body aches, loss of taste or smell, fever, chills, and fatigue, *etc*.; (4) followed by patients with concerns for various infections.

Asymptomatic patients were categorized by finding a COVID positive test, ruling out any URI. If a patient was not admitted to the hospital with URI symptoms, the chief complaint was not COVID related, and there was a lack of symptoms, this patient was counted as asymptomatic.

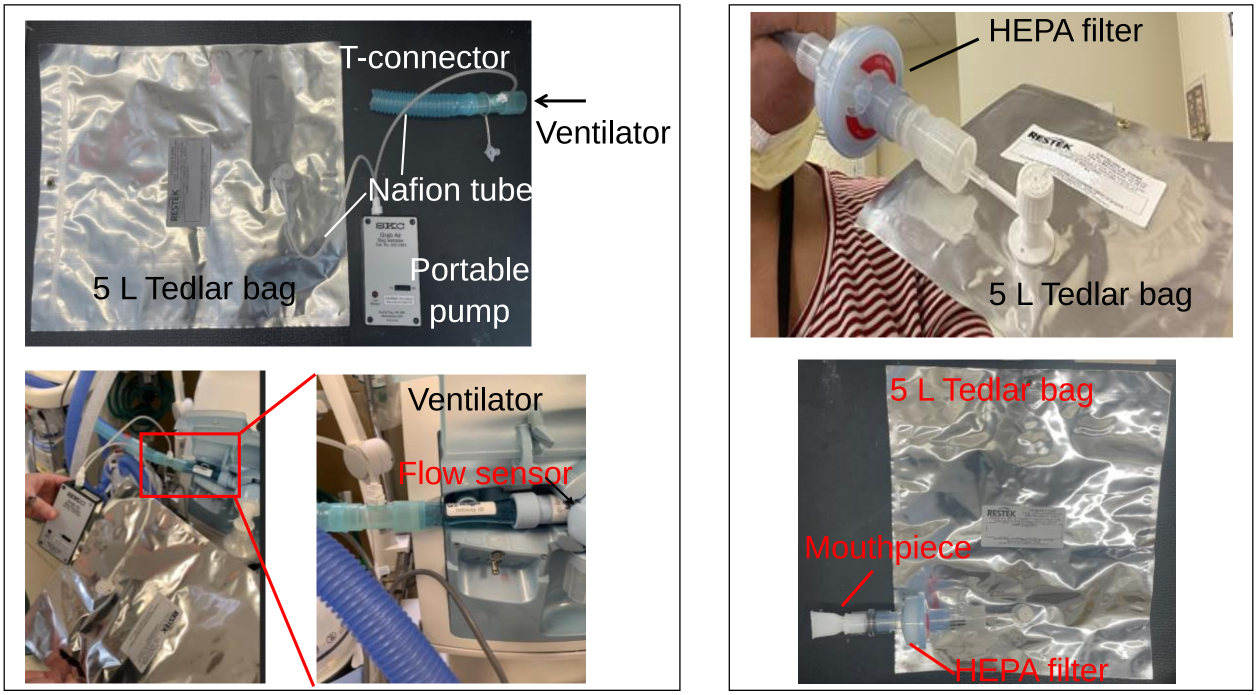


**Figure S1. (Left panel)** Breath collection from a ventilated patient. A portable pump was used to pump breath into a 5 L Tedlar bag via a T-connector connected to the flow sensor on the ventilator expiratory port. **(Right panel)** Breath collection from a non-ventilated patient into a 5 L Tedlar bag via a mouthpiece and an in-line HEPA filter.

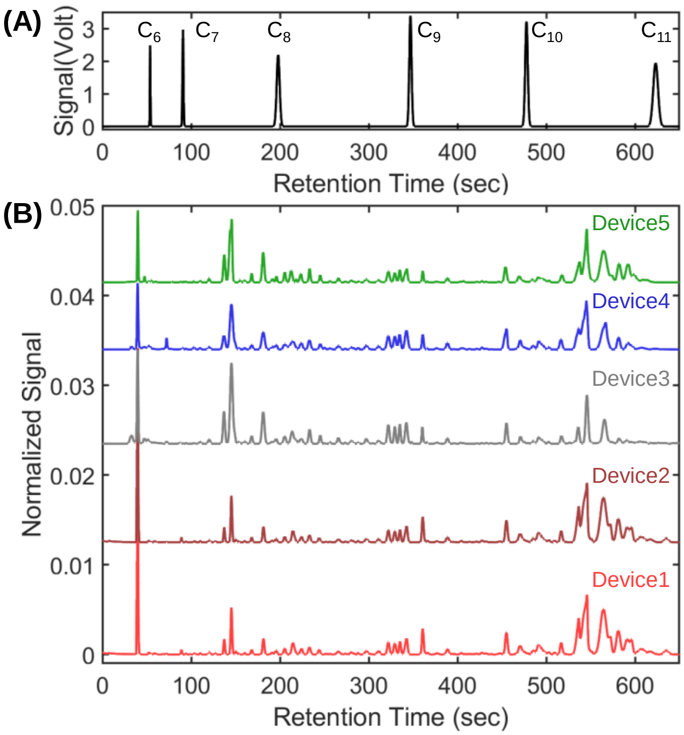


**Figure S2.** Inter-GC characterization of five portable GC devices used in the study with the same breath sample collected from a non-COVID subject. Note that the chromatogram of $C_6$-$C_{11}$ **(A)** obtained from one of the five GC devices operated under identical conditions (such as temperature ramping, and flow rate, *etc*.) is superimposed on those human breath chromatograms **(B)** to mark the breath VOC peaks against those of $C_6$-$C_{11}$.

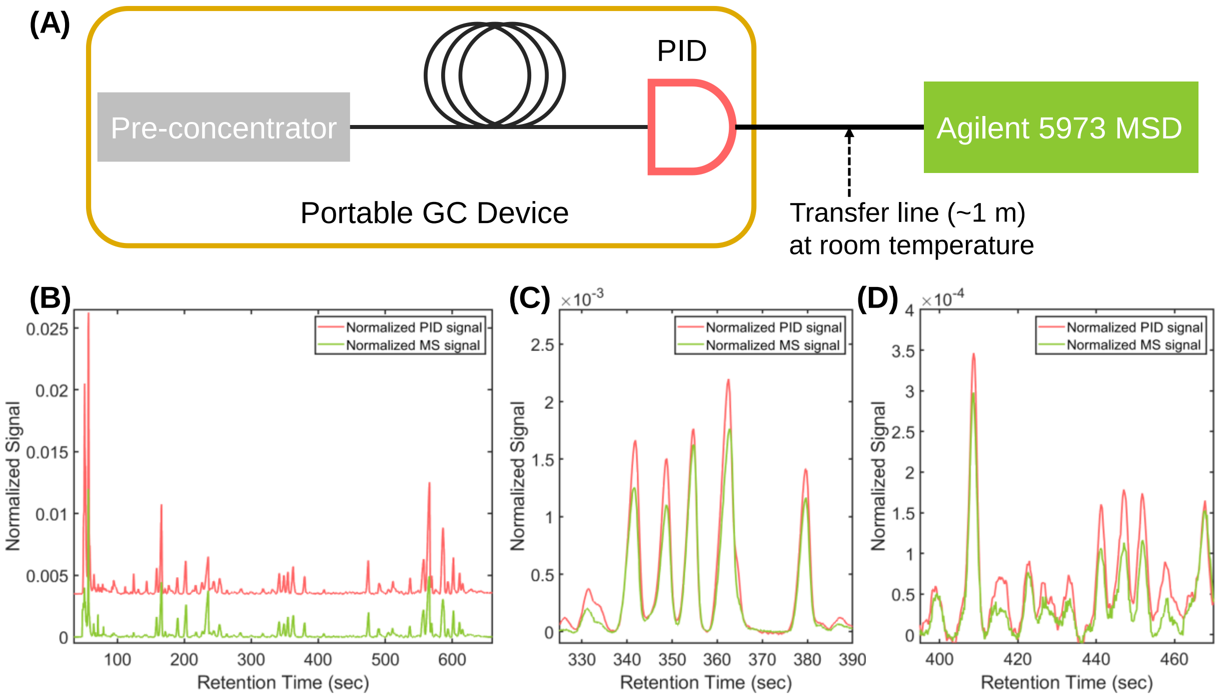


**Figure S3. (A)** Setup that connects the outlet of our portable GC to an Agilent mass spectrometry (MS). **(B)** Chromatograms obtained concomitantly by the photoionization detector (PID) in our GC and MS for the same breath sample. For easy comparison, both chromatograms are normalized to the total area under the curve (from 0 s to 650 s). **(C)** and **(D)** Two zoom-in portions in (B).

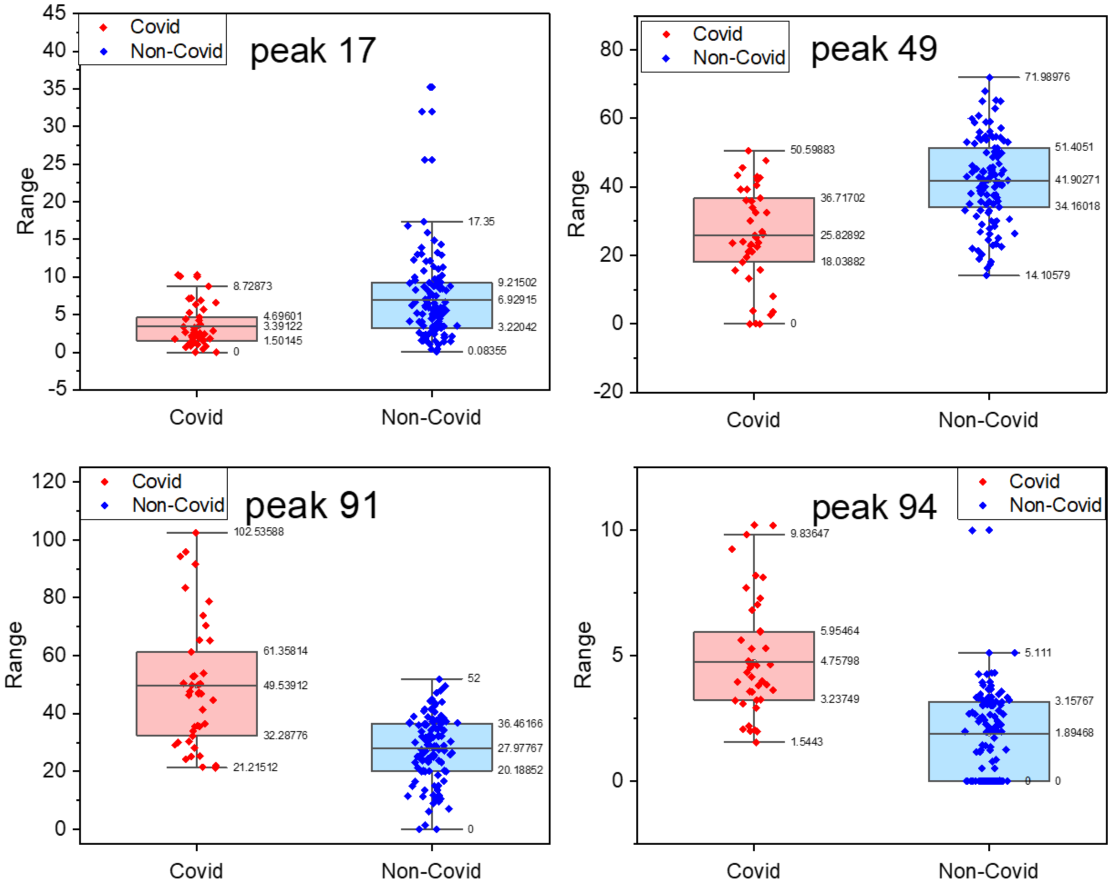


**Figure S4.** Violin chart of 4 biomarkers used in Figure 3 to distinguish between COVID (2021) and non-COVID (all COVID negative recruited throughout the study).

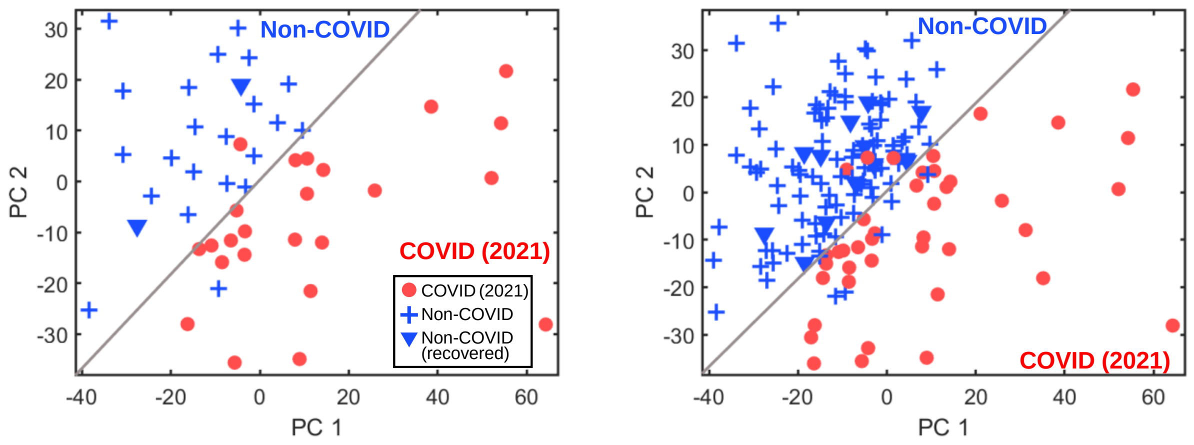


**Figure S5.** PCA plot using the 4 VOC biomarkers in Figure 3 and Table 1 to distinguish between COVID (2021) and non-COVID. **(A)** Training set. **(B)** Training set plus testing set. The corresponding statistics are given in Table 2. COVID (2021) patients (red circles) refer to those who were recruited prior to December 14, 2021, and were therefore assumed to be infected by Delta or earlier variants. Non-COVID patients (blue crosses) refer to those who were recruited throughout the study (from April 26, 2021 to May 31, 2022). They were COVID negative at the time when breath analysis was conducted and their COVID infection history was unknown. Non-COVID (recovered) patients (blue triangles) refer to those who were recruited throughout the study and had previously been COVID positive, but recovered, *i.e.*, COVID negative at the time of breath analysis was conducted. Each data point in the PCA plot represents one different breath sample. The breath sample was obtained and analyzed within 18 hours of the PCR test. The gray line marks the boundary of COVID and non-COVID. The bottom right zone represents the COVID region, whereas top left zone represents the non-COVID region.

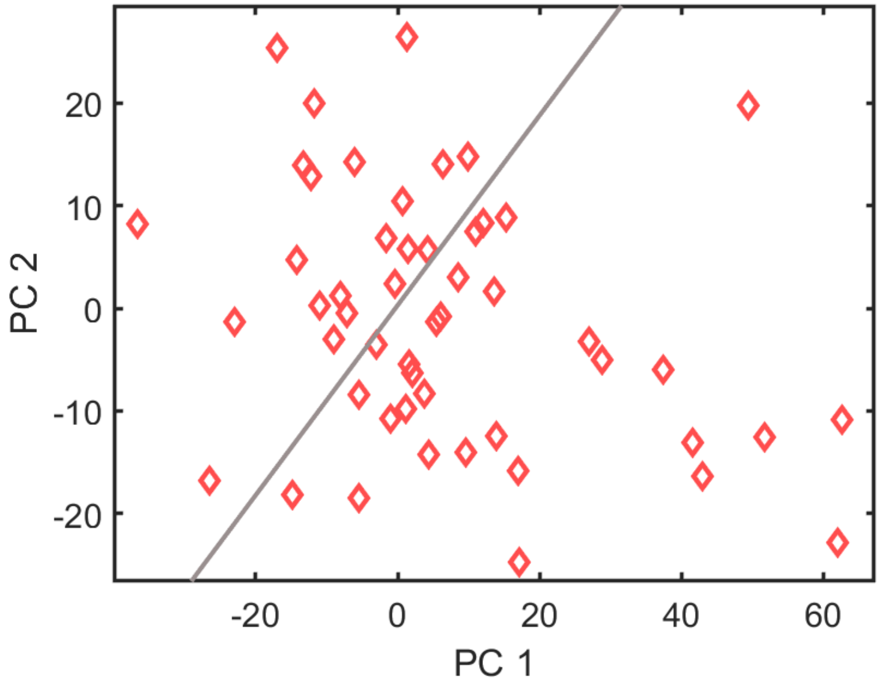


**Figure S6**. PCA plot for COVID (2022) patients when the 4 VOC biomarkers in Figure 3 for COVID (2021) are used. A significantly lower specificity, sensitivity, and accuracy are obtained, which suggests that we cannot use the same set of biomarkers for Delta (and earlier variants) for Omicron. COVID (2022) patients refer to those who were recruited after January 11, 2022 (till the end of the study – May 31, 2022) and were therefore assumed to be infected by Omicron. COVID (2021) patients refer to those who were recruited between prior to December 14, 2021, and were therefore assumed to be infected by Delta and earlier variants.

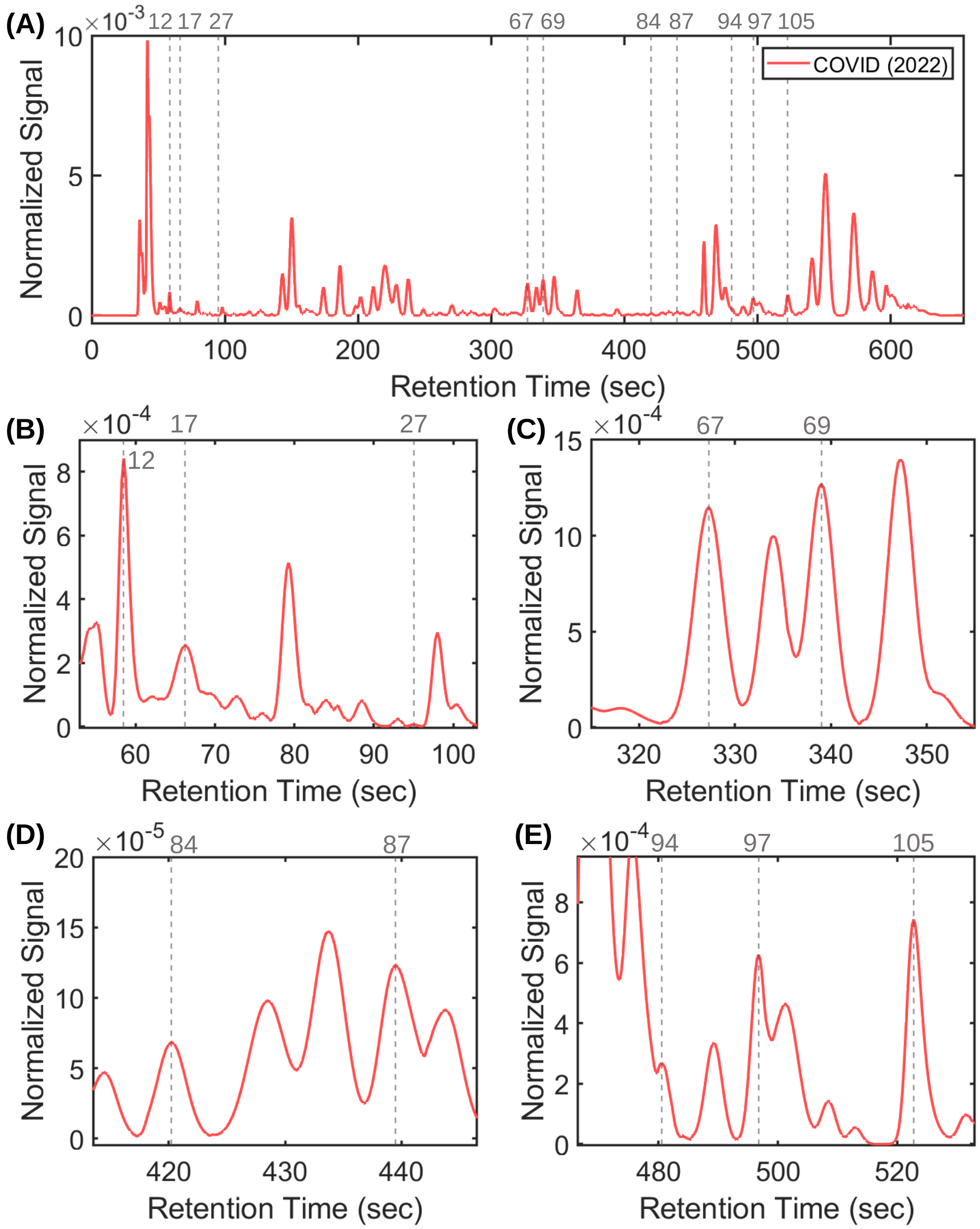


**Figure S7.** Positions of the biomarkers used in Figure S8 to distinguish between COVID (2022) and non-COVID (Peak ID: 17, 67, 87, 97), in Figure S9 to distinguish between COVID (all variants occurring between April 2021 and May 2022) and non-COVID (Peak ID: 12, 67, 84, 105), and in Figure S10 to distinguish between COVID (2021) and COVID (2022) (Peak ID: 27, 67, 69, 87, 94). The names of those VOCs are given in Table 1.

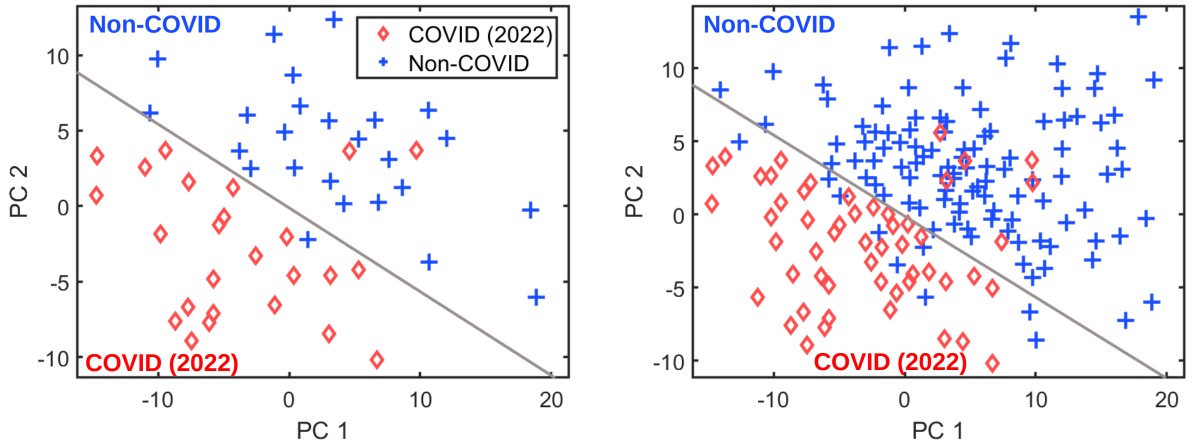


**Figure S8.** PCA plot using 4 VOC biomarkers in Figure S7 and Table 1 to distinguish between COVID (2022) and non-COVID. **(A)** Training set. **(B)** Training set plus testing set. The corresponding statistics are given in Table 3. COVID (2022) patients (red diamonds) refer to those who were recruited after January 11, 2022 (till the end of the study – May 31, 2022) and were therefore assumed to be infected by Omicron. Non-COVID patients (blue crosses) refer to those who were recruited throughout the study (from April 26, 2021 to May 31, 2022). They were COVID negative at the time when breath analysis was conducted and their COVID infection history was unknown, or recovered (*i.e.*, previously COVID positive, but COVID negative at the time when breath analysis was conducted). Each data point in the PCA plot represents one different breath sample. The breath sample was obtained and analyzed within 18 hours of the PCR test. The gray line marks the boundary of COVID and non-COVID. The bottom left zone represents the COVID region, whereas the top right zone represents the non-COVID region.

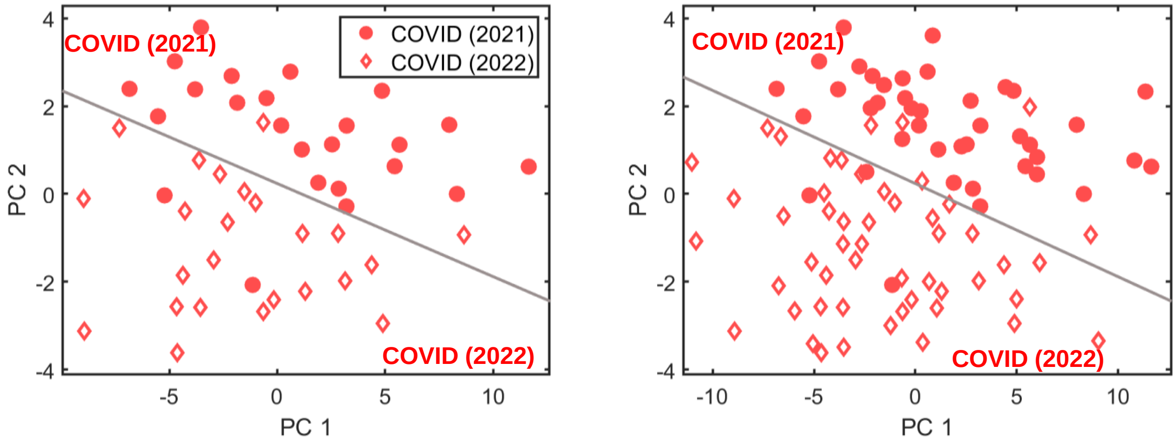


**Figure S9.** PCA plot using 5 VOC biomarkers in Figure S7 and Table 1 to distinguish between Omicron and the previous variants. **(A)** Training set. **(B)** Training set plus testing set. The corresponding statistics are given in Table 4. Omicron patients, COVID (2022), are denoted as red diamonds. They are the same as presented in Figures S8. All previous variants' patients, COVID (2021), are denoted as red circles. They are the same as presented in Figures S5. Each data point in the PCA plot represents one different breath sample. The breath sample was obtained and analyzed within 18 hours of the PCR test. The gray line marks the boundary of Omicron and previous variants. The bottom left zone represents the Omicron region, whereas the top right zone represents the previous variants region.

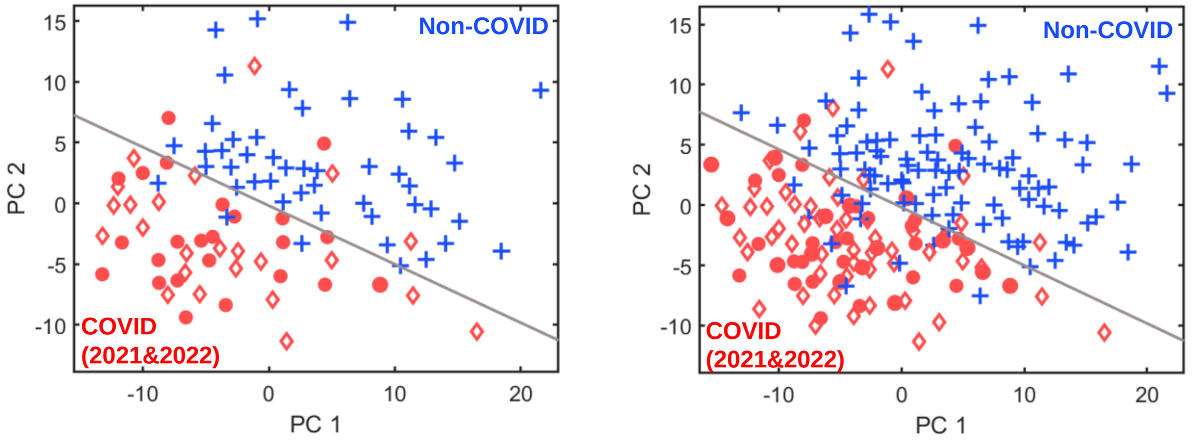


**Figure S10.** PCA plot using 4 VOC biomarkers in Figure S7 and Table 1 to distinguish between COVID (all variants) and non-COVID. **(A)** Training set. **(B)** Training set plus testing set. The corresponding statistics are given in Table 5. COVID patients are denoted as red circles for COVID (2021) and red diamonds for COVID (2022), respectively. All non-COVID patients are denoted as blue crosses. All the COVID and non-COVID (including the recovered) patients were recruited throughout the study from April 26, 2021 to May 31, 2022. Each data point in the PCA plot represents one different breath sample. The breath sample was obtained and analyzed within 18 hours of the PCR test. The gray line marks the boundary of COVID and non-COVID. The bottom left zone represents the COVID region, whereas the top right zone represents the non-COVID region.

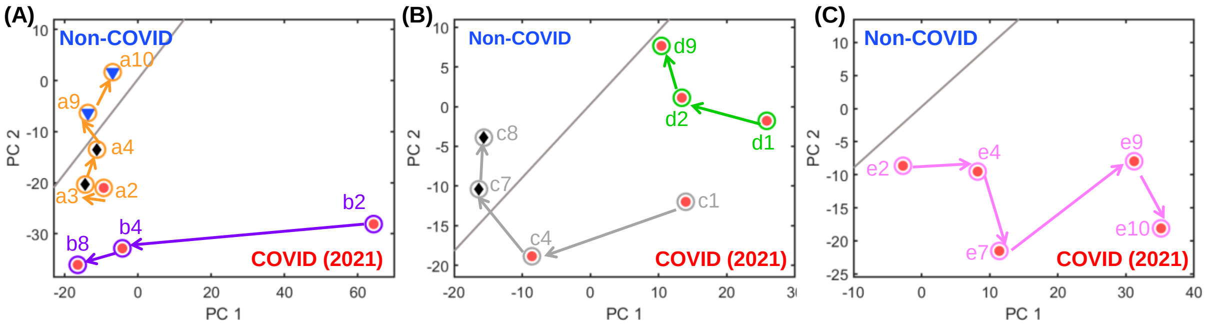


**Figure S11.** Trajectories on the PCA plot for various patients monitored for multiple days. **(A)** and **(B)** are the recovery cases. The patients were initially COVID positive, and later recovered and discharged from the hospital. Patient *b* on Day 2 (data point marked as *b*2) represents the most severe case that we saw among the entire patient pool, whose data point is the farthest in distance from the boundary (grey line). **(C)** A deterioration case. This patient (Patient *e*) was COVID positive and his/her situation deteriorated over time. This patient died 21 days after our last GC measurement (Day 10). All the patients were recruited prior to December 14, 2021, and were therefore assumed to be infected by Delta or earlier variants. Each data point in the figure is denoted as "LowerCaseLetterNumber". For example, *b*2 refers to the data point of Patient *b*, whose breath collection/analysis was conducted on Day 2 after this patient was recruited into our study. Red circles and blue triangles denote COVID positive and negative (recovered), respectively, which were confirmed by the PCR tests within 18 hours of breath collection/analysis. These data points also appear in the PCA plots in Figure S5, and are tallied in Figure 1 and statistics in Table 2. Black diamonds denote the breath data points that the corresponding PCR tests within 18 hours were not available. These data points are not plotted in Figure S5, nor are they used/tallied in Figure 1 or Table 2.

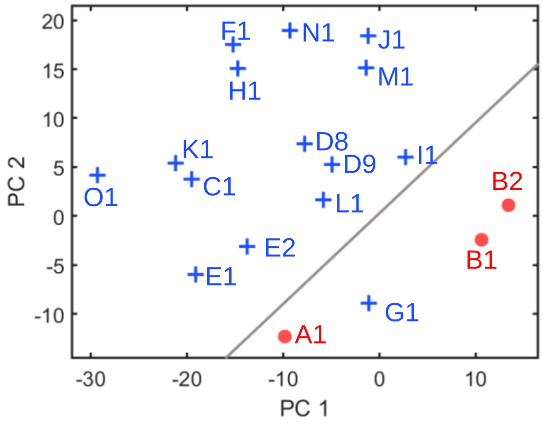


| Patient | Patient ID | Virus type |
|---|---|---|
| A | 121321A | COVID (2021), asymptomatic |
| B | 100521B | COVID (2021), asymptomatic |
| C | 072121A | HCoV-OC43 |
| D | 081021B | Rhinovirus |
| E | 121421A | Rhinovirus |
| F | 020822A | Human metapneumo virus |
| G | 021022B | Parainfluenza virus 3 |
| H | 031022A | Rhinovirus |
| I | 033022A | HCoV-OC43 |
| J | 033122B | Rhinovirus& Enterovirus |
| K | 040122B | Parainfluenza virus 3 |
| L | 040522A | Rhinovirus&enterovirus, influenza A |
| M | 041222B | Rhinovirus |
| N | 042722A | HCoV-OC43 |
| O | 050922D | Rhinovirus |

**Figure S12** Asymptomatic COVID (2021) patients and patients who were infected by other viruses on the same PCA plot as in Figure S5. All COVID positive patients (red circles) were recruited prior to December 14, 2021, and were therefore assumed to be infected by Delta or earlier variants. All non-COVID patients (blue crosses) were recruited throughout the study (from April 26, 2021 to May 31, 2022). Each data point in the figure is denoted as “UpperCaseLetterNumber”. For example, B2 refers to the data point of Patient B, whose breath collection/analysis was conducted on Day 2 after recruitment. The COVID status of each data point was confirmed by the PCR test within 18 hours of breath collection/analysis. These data points have appeared in the PCA plots in Figure S5 and tallied in Figure 1 and statistics in Table 2. The types of viruses are listed in the table above.

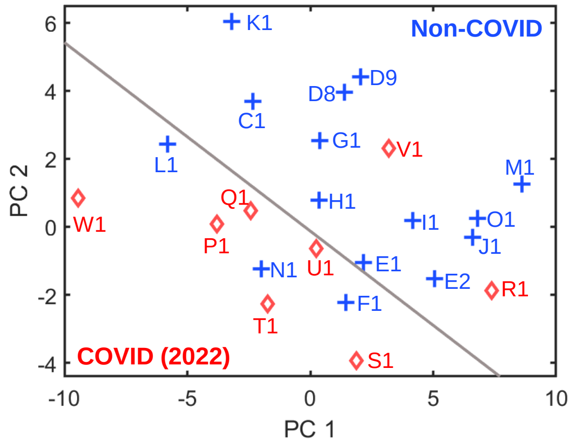


| Patient | Patient ID | Virus type |
|---|---|---|
| P | 020822B | COVID (2022), asymptomatic |
| Q | 022222A | COVID (2022), asymptomatic |
| R | 042522A | COVID (2022), asymptomatic |
| S | 042822D | COVID (2022), asymptomatic |
| T | 050922A | COVID (2022), asymptomatic |
| U | 051222D | COVID (2022), asymptomatic |
| V | 051622B | COVID (2022), asymptomatic |
| W | 051622D | COVID (2022), asymptomatic |
| C | 072121A | HCoV-OC43 |
| D | 081021B | Rhinovirus |
| E | 121421A | Rhinovirus |
| F | 020822A | Human metapneumo virus |
| G | 021022B | Parainfluenza virus 3 |
| H | 031022A | Human Rhinovirus |
| I | 033022A | HCoV-OC43 |
| J | 033122B | Rhinovirus& Enterovirus |
| K | 040122B | Parainfluenza virus 3 |
| L | 040522A | Rhinovirus&enterovirus, influenza A |
| M | 041222B | Rhinovirus |
| N | 042722A | HCoV-OC43 |
| O | 050922D | Rhinovirus |

**Figure S13.** Asymptomatic COVID (2022) patients and patients who were infected by other viruses on the same PCA plot as in Figure S8. All COVID positive patients (red diamonds) were recruited after January 11, 2022, and were therefore assumed to be infected by Omicron. All non-COVID patients (blue crosses) were recruited throughout the study (from April 26, 2021 to May 31, 2022). Each data point in the figure is denoted as "UpperCaseLetterNumber", as in Figure S12. The COVID status of each data point was confirmed by the PCR test within 18 hours of breath collection/analysis. These data points have appeared in the PCA plots in Figure S8 and tallied in Figure 1 and statistics in Table 3. The types of viruses are listed in the table above.

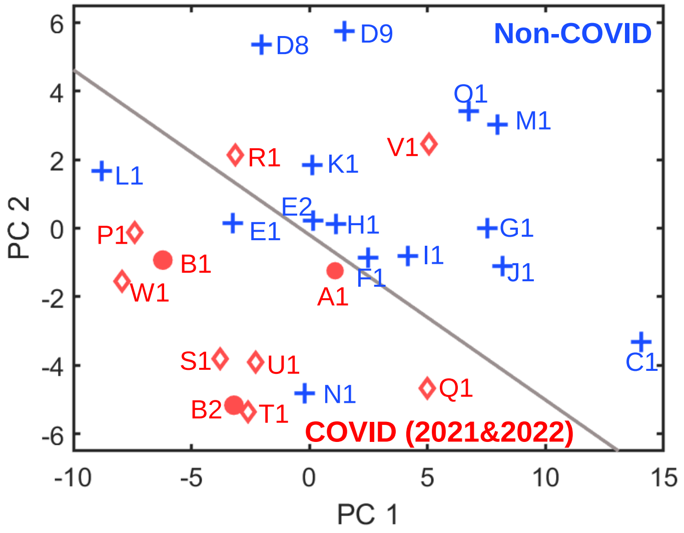


**Figure S14.** Asymptomatic COVID patients (regardless of variants, red dots and diamonds for COVID (2021) and COVID (2022), respectively) and non-COVID patients who were infected by other viruses (blue crosses) on the same PCA plot as in Figure S9. Each data point in the figure is denoted as "UpperCaseLetterNumber", as in Figure S12. The COVID status of each data point was confirmed by the PCR test within 18 hours of breath collection/analysis. These data points have appeared in the PCA plots in Figure S9 and tallied in Figure 1 and statistics in Table 4. The types of viruses are listed in the table above.
